# Cluster Analysis of Allergic Poly-Sensitizations in Urban Adults with Asthma

**DOI:** 10.1101/2022.05.08.22274727

**Authors:** Brian J. Patchett, Bede N. Nriagu, Granit Mavraj, Ruchi R. Patel, Christopher MacLellan, Tushar Thakur, Edward S. Schulman

## Abstract

**Introduction:** While reliable, quantitative *in vitro* testing for sensitivity to aeroallergens has been available for decades, if and how asthma severity markers might be predictably expressed in clusters matched for comparable multiple sensitizations is unknown.

**Objective:** Our aim is to use machine learning techniques to explore how allergic poly-sensitization (APS) clusters may serve as precision markers in adult urban patients with moderate to severe asthma.

**Methods:** We constructed a database of sensitizations to the 25 aeroallergens in the Zone 1 Northeastern US ImmunoCAP® assay. We used the Scikit-Learn® machine learning library to perform model-based clustering to identify APS clusters. Clusters were compared for differences in common clinical markers of asthma.

**Results:** The database consisted of 509 patients. Unbiased mixture modeling identified ten clusters of increasing APS of varying size (n = 1 to 339) characterized by significant increases in mean serum immunoglobulin E (p<.001), peripheral blood eosinophil count (p<.001), and D_LCO_ (p=.02). There was a significant decline in mean age at presentation (p<.001), FEV_1_/FVC (p=.01), and FEF_25-75_ (p=.002), but not FEV_1_ (p=.29), nor RV/TLC (p=.14) with increasing APS by simple linear regression. Finally, we identified two divergent paths for the poly-atopic march, one driven by perennial and the other by seasonal allergens.

**Conclusion:** We conducted a pilot study for a novel machine learning understanding and approach to the classification of APS and potential influences if included in asthma cluster analyses. The methods used here can be easily applied to other geographic regions with different allergens

## INTRODUCTION

Asthma is known as both one of the most common chronic respiratory conditions as well as a highly heterogeneous disease^1^. An active area of research has been characterizing the heterogeneity of asthma and assessing how cohorts with similar characterizations can predict more precision treatments^2,3^. In recent years, machine learning-derived cluster analyses have identified subgroups that share asthma phenotypes and endotypes. Haldar et al^4^ from the UK was one of the first groups to identify severe asthma clusters and would be soon followed by the Severe Asthma Research Program^5^ (SARP) from the NIH/NHLBI. As research groups explore findings across large and diverse cohorts^6-8^, a goal is to prospectively identify markers useful to classify patients within well characterized clusters^9-13^. Atopic clusters, identified by the UK, US and other independent analyses,^14,15^ are of particular interest for their high disease burden and medication usage.

In atopic asthma, the Immunoglobulin E (IgE)-mediated sensitization to an allergen(s) and subsequent re-exposure is a major driver of asthmatic symptoms and exacerbations^16,17^. Between 51 and 80% of patients will have sensitization to more than one allergen, a state referred to as poly-sensitization^18^. Moreover, poly-sensitized patients will experience more severe allergy and asthma symptoms than mono-sensitized patients^19^. To-date however, cluster analyses have largely disregarded laboratory quantitation of atopic sensitization, used self-reported atopy or included atopy as a touchpoint in their analysis if one or more skin prick tests was positive over control (Table 1). These analyses have not considered the number of sensitizations, quantitative level of sensitization as measured by allergen specific IgE, patterns of co-sensitization, or synergy between specific allergens as either variables in the clustering or as a biomarker to predict cluster assignment. Thus, an individual with one weakly positive skin test or IgE-allergen sensitization and an individual that is highly poly-sensitized are both labeled equally atopic. The focus of this study is the number of sensitizations and clustering of specific IgE levels in the aeroallergen panel^20,21^ from patients in our urban asthma clinic. Moreover, we assess how demographic and phenotypic markers of asthma might be predictably influenced in patients matched for similar positive allergens and levels of sensitivity to those allergens.

**TABLE 1:**
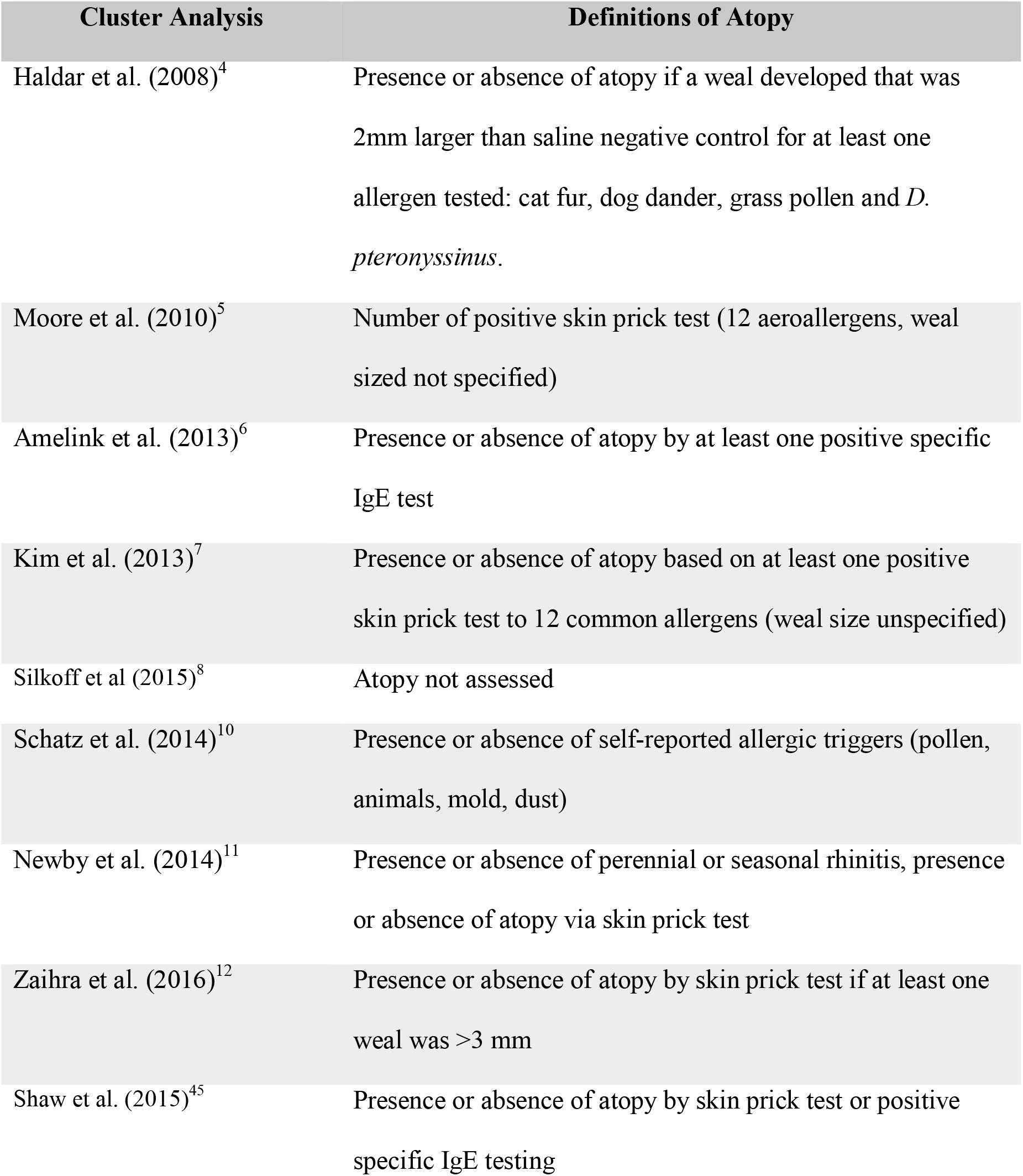
Major severe asthma cluster analyses and how atopy was defined.

| Cluster Analysis | Definitions of Atopy |
| --- | --- |
| Haldar et al. (2008)<sup>4</sup> | Presence or absence of atopy if a weal developed that was 2mm larger than saline negative control for at least one allergen tested: cat fur, dog dander, grass pollen and *D. pteronyssinus*. |
| Moore et al. (2010)<sup>5</sup> | Number of positive skin prick test (12 aeroallergens, weal sized not specified) |
| Amelink et al. (2013)<sup>6</sup> | Presence or absence of atopy by at least one positive specific IgE test |
| Kim et al. (2013)<sup>7</sup> | Presence or absence of atopy based on at least one positive skin prick test to 12 common allergens (weal size unspecified) |
| Silkoff et al (2015)<sup>8</sup> | Atopy not assessed |
| Schatz et al. (2014)<sup>10</sup> | Presence or absence of self-reported allergic triggers (pollen, animals, mold, dust) |
| Newby et al. (2014)<sup>11</sup> | Presence or absence of perennial or seasonal rhinitis, presence or absence of atopy via skin prick test |
| Zaihra et al. (2016)<sup>12</sup> | Presence or absence of atopy by skin prick test if at least one weal was >3 mm |
| Shaw et al. (2015)<sup>45</sup> | Presence or absence of atopy by skin prick test or positive specific IgE testing |

## METHODS

### Chart Review

We performed a retrospective chart review of adults with moderate to severe asthma who presented to the Drexel University outpatient Pulmonary/Asthma Clinic between 2008 to 2019 and analyzed IgE levels for each positive allergen in the Zone 1 Northeastern US ImmunoCAP® 25 aeroallergen panel (see supplementary tabble 1 for list of allergens). During the study period, our asthma clinics exclusively ordered the widely commercially available ImmunoCAP® assays for identification of allergen sensitizations. All tests were performed by either Labcorp or Quest Diagnostics. Multiple studies have shown this assay to be quantitative, accurate and reproducible^20-22^. Though we ordered the Zone 1 profile, specific allergen profiles are available for twenty different US regions including Puerto Rico. The research was approved by the Drexel Institutional Review Board in an expedited review. Consent was not required because of de-identification. Diagnosis of moderate or severe asthma was made consistent with Step 4 to 6 GINA and/or ERS/ATS guidelines^23, 24^. Information extracted included self-identified demographics, smoking history, BMI, asthma and allergy medications, comorbid conditions (e.g., COPD, rhinitis), total IgE level, pulmonary function tests (PFT), and earliest recorded peripheral blood eosinophil count (PBEC). All extracted information came from or was ordered at the patient’s initial specialist visit. Missing data were not imputed. Our exclusion criteria included patients <18 years old, those receiving allergen-specific immunotherapies, systemic corticosteroid use within a month of blood draw, hematologic malignancies, and allergic bronchopulmonary aspergillosis. Patients with asthma-COPD overlap (ACO) were included if there was a history of childhood asthma or physician documented asthma before age 30. A high poverty zip code was defined as having a poverty rate higher than the national average, 10.5%^26^, from the City of Philadelphia Health Dashboard^27^.

### Preprocessing

During the study period the ImmunoCAP® lower detection limit changed from “<0.35 IU/ml” to “<0.10 IU/ml” from certain laboratory providers. In either case, when the test reported less than the lower limit, we entered a “0” into our database. If the upper limit of detection (>100 IU/ml) was reported, we entered “100” into our database. Though normalization is a common step in machine learning data processing, because the sensitization data was on the same scale (i.e., all variables were specific IgE with the same range) this was not necessary and therefore was not done. We used the elbow method with a scree plot^28^ (supp. fiig 1) for dimensionality reduction; the elbow occurred between 3 and 6 principal components. We selected 4 principal components because the resulting clustering best balanced explaining the data without overfitting.

### Dirichlet Process Mixture Models

To identify APS profiles, we performed cluster analysis in the form of Dirichlet Process Mixture Modeling (DPMM)^29^ on the ImmunoCAP® 25 aeroallergen specific IgE panel by utilizing the Scikit-Learn machine learning library^30^ in Python version 3.9.0. This library uses a Variational Bayes (VB)^31, 32^ approach with a Gaussian process prior to determine the cluster number, means, and covariance matrices as well as cluster assignment. Unlike other clustering algorithms where the user must predetermine the appropriate cluster number, with a VB approach the user is only required to input an upper bound for the appropriate cluster number. With this upper bound, some of the clusters will be used while others will be given a weight of zero and contain no observations/patients^33^. In this work, a cluster number of 20 was estimated based on preliminary clustering trials to be a valid upper limit. All other parameters required for DPMMs were kept at the Scikit-Learn program-calculated defaults.

### Assessment of Cluster Correlations

After visualizing the clustering results, it was hypothesized sensitization to each allergen did not increase uniformly between clusters. A correlation circle was plotted with the contribution of each allergen to the first two principal components^34^. Viewing these findings, we further hypothesized there would exist clusters of the clusters, here called “superclusters”. To evaluate this, we created a correlation matrix of the clusters, then ordered the matrix by hierarchical clustering. Correlation matrix plotting was done with corrplot^35^, and factor analysis was done with the factoextra^36^ and FactoMineR^37^ packages in RStudio version 1.3^38^.

### Between Cluster Comparison of Clinical Markers

Given the known relationship between IgE and asthma, we hypothesized clusters associated with higher levels of sensitization would be predictive of more perturbed asthmatic clinical markers. To assess this, we first calculated an APS score for each cluster defined by the overall average allergen sensitization. Simple linear regression was then used to assess the relationship between the cluster APS scores and common clinical markers of allergic asthma, including serum IgE, PBEC, PFT results, and demographic data. The data for serum total IgE and PBEC were linearized prior to regression analysis. Plotting was done with ggplot2^39^ and dplyr^40^.

## RESULTS

### Chart Review

A demographic review of the 509 patients who met inclusion criteria is shown in Table 2. The predominant racial group was Black/African American (61.9%). Most patients identified as female (77.1%). The predominant age range was 36-64 years old (mean = 50.4 ± 13.9, range of 20 to 88 years). BMI average was 33.0 ± 9.3 kg/m^2^ (range of 16.3 to 72.1 kg/m^2^). A large majority (76.2%) resided in high poverty zip codes. This was particularly evident among Black (87.8%) and Latino (88.3%) patients. Current and past smoking history of ≥10 pack/years was documented in 40.1% of patients.

**TABLE 2:** Demographics of subjects in the study: †only the latter 179 patients had their gender recorded.

| **Demographics of subjects** |  |
| --- | --- |
| **N** | 509 |
| **Age (SD)** | 50.48 (13.9) |
| **Age Groups (%)** |  |
| 18-35 years | 84 (16.6) |
| 36-64 years | 349 (69.0) |
| ≥ 65 years | 73 (14.4) |
| Missing | 3 (0.58) |
| **Male:Female Ratio†** | 1:3.4 |
| Percent Male, Percent Female | 22.9%, 77.1% |
| **BMI kg/m<sup>2</sup> (SD)** | 33.0 (9.3) |
| **BMI Groups (%)** |  |
| < 18.5kg/m<sup>2</sup> (%) | 12 (2.4) |
| 18.5 – 24.9kg/m<sup>2</sup> (%) | 89 (17.5) |
| 25.0 – 29.9kg/m<sup>2</sup> (%) | 93 (18.2) |
| ≥ 30kg/m<sup>2</sup> (%) | 267 (52.4) |
| Missing | 48 (9.4) |
| **Race (%)** |  |
| Black | 315 (61.9) |
| White | 131 (25.7) |
| Latino | 26 (5.1) |
| Asian | 23 (4.5) |
| Other/Not specified | 14 (2.8) |
| **In zip codes of high poverty (%)** | 76.2% |
| Black | 270 (87.8) |
| White | 61 (48.0) |
| Latino | 22 (88.3) |
| Asian | 18 (78.3) |
| Other/Not specified | 3 (50) |
| **Smoking ≥ 10 pack/years (%)** | 198/494 (40.1) |

### Ten Component Model for APS

DPMMs generated 10 clusters with a non-zero weight, and the 10 clusters were given a weight of zero (Figure 2a). The demographic makeup including number of patients with missing data within clusters is shown in Table 3. The largest cluster was cluster one (n=339). These patients averaged 52.3 ± 13.9 years old, had an average BMI of 33.0 ± 9.5 kg/m^2^, and most (73.3%) lived in high poverty zip codes. Cluster two consisted of 61 patients who tended to be slightly younger at 46.9 ± 12.9 years old. There were 51 patients in cluster three and had a similar age at 48.9 ± 13.7 years old. Clusters four through ten had 25 or fewer patients each. Though, obviously representing rare patients, the unbiased VB algorithm exposed separate, distinct, and high-level polysensitization patterns in the four patients in clusters eight through ten (see below).

**TABLE 3:** Demographics of each cluster. Mean and standard deviation are given where appropriate.

| **Demographic Data** | **Cluster 1** | **Cluster 2** | **Cluster 3** | **Cluster 4** | **Cluster 5** | **Cluster 6** | **Cluster 7** | **Cluster 8** | **Cluster 9** | **Cluster 10** |
| --- | --- | --- | --- | --- | --- | --- | --- | --- | --- | --- |
| Total n | 339 | 60 | 51 | 25 | 6 | 16 | 8 | 2 | 1 | 1 |
| Age (SD) | 52.3<br>(13.9) | 46.9<br>(12.9) | 48.9<br>(13.7) | 48.7<br>(11.0) | 52.8<br>(9.7) | 45.8<br>(12.4) | 32.6<br>(14.3) | 30<br>(8.5) | 43 | 21 |
| Missing, n (%) | 1 (0.3) | 0 (0) | 1 (4.2) | 0 (0) | 0 (0) | 0 (0) | 0 (0) | 0 | 0 | 0 |
| BMI (SD) | 33.0<br>(9.5) | 33.9<br>(9.1) | 34.2<br>(9.1) | 30.4<br>(8.5) | 33.6<br>(7.6) | 33.3<br>(10.7) | 29.5<br>(7.5) | 31.1<br>(4.8) | 28.0 | 21.7 |
| Missing, n (%) | 28 (8.9) | 13 (18.1) | 2 (8.3) | 5 (14.3) | 1 (8.3) | 0 (0) | 1 (10) | 0 (0) | 0 (0) | 0 (0) |
| High poverty zip code, n (%) | 241<br>(73.3) | 46<br>(78.0) | 42<br>(82.4) | 23<br>(100) | 3<br>(60) | 13<br>(81.6) | 8<br>(100) | 2<br>(100) | 0<br>(0) | 0<br>(0) |
| Missing, n (%) | 10 (2.9) | 1 (1.7) | 0 (0) | 2 (8.0) | 1 (16.7) | 0 (0) | 0 (0) | 0 (0) | 0 (0) | 0 (0) |
| Race |  |  |  |  |  |  |  |  |  |  |
| Black | 190 | 43 | 40 | 20 | 1 | 12 | 6 | 1 | 1 | 1 |
| White | 100 | 14 | 6 | 3 | 4 | 3 | 0 | 1 | 0 | 0 |
| Latino | 21 | 2 | 0 | 2 | 1 | 0 | 0 | 0 | 0 | 0 |
| Asian | 19 | 0 | 2 | 0 | 0 | 0 | 2 | 0 | 0 | 0 |
| Other | 9 | 1 | 3 | 0 | 0 | 1 | 0 | 0 | 0 | 0 |

The spread of the sensitizations for each cluster was right-skewed, therefore the mean and standard deviation (Table 4a-c) as well as the median and interquartile range (Table 4d-f) are reported. Broadly, cluster assignment was determined by both which allergens the patient was sensitive to and their specific IgE levels. Assignment to cluster 1 appeared to be driven by a median of one positive sensitization (range zero to three, see supplementary tabble 2). The APS score was only 0.11 IU/ml. The overall mean specific IgE increased with respect to cluster number, but not every allergen increased stepwise with each cluster. Cluster 2 patients had a APS of 1.39 IU/ml with notably high sensitizations to dog and especially cat. Cluster 3 had a similar APS to cluster 2 of 1.69 IU/ml and tended to have high sensitization to *D. pteronyssinus* and *D. farinae* with comparatively modest sensitizations to the other allergens. Cluster 4 had a higher APS of 4.25 IU/ml with high sensitizations to Timothy and Bermuda grass, birch, oak, maple trees, as well as cat and dog. Cluster 5 had an APS of 7.07 IU/ml. Assignment appeared driven by high-level sensitizations to cat and dog, as well as a general increase in all seasonal allergies but low sensitization to molds. Similar to cluster 5, cluster 6 had a APS of 7.35 IU/ml, driven by high-level sensitizations to *D. pteronyssinus*, and *D. farinae*, as well as cat and dog. The APS of cluster 7 was 13.55 IU/ml, exposing prominent sensitizations to birch and oak tree, as well as cat and dog.

**TABLE 4:**
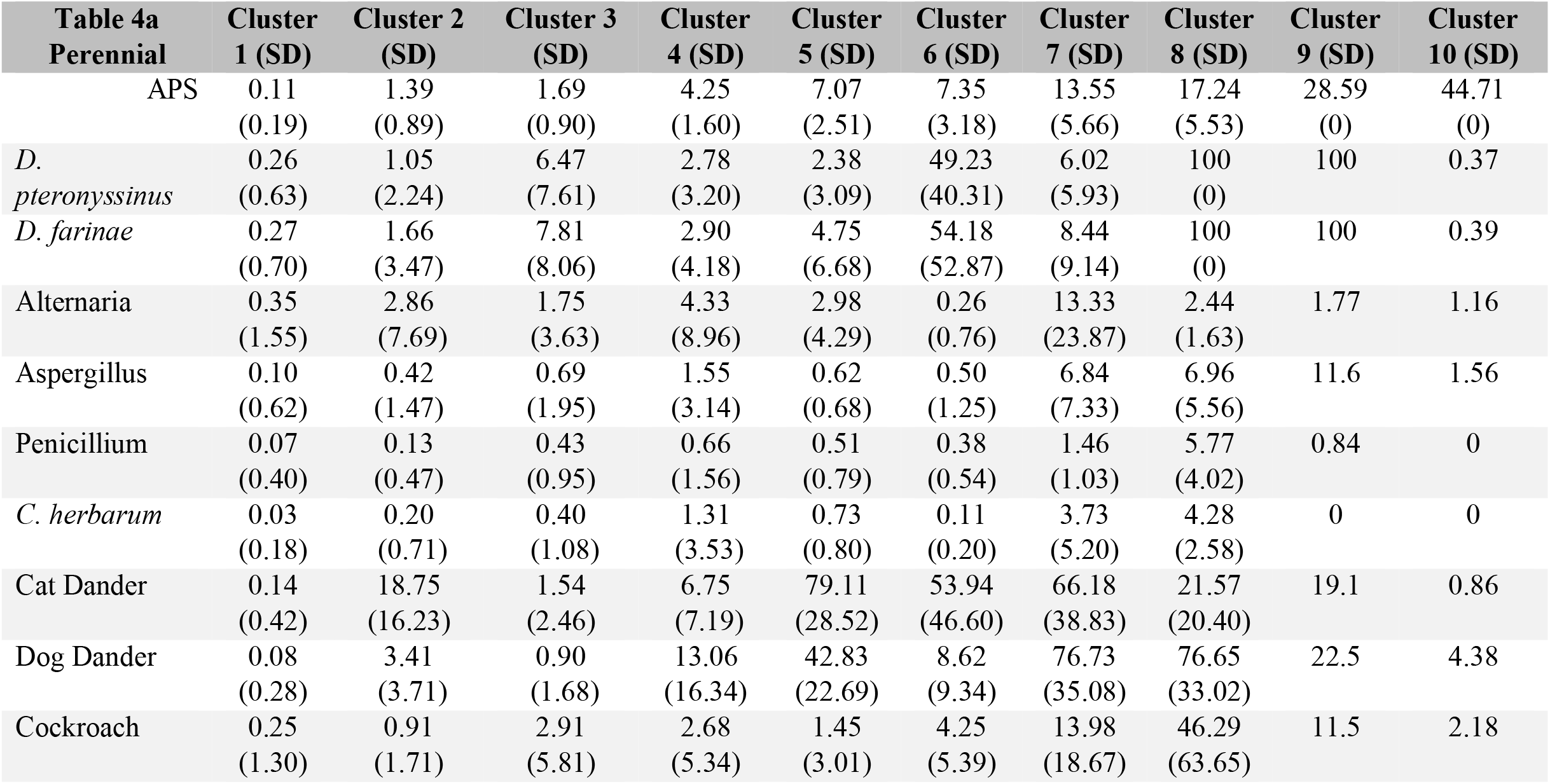
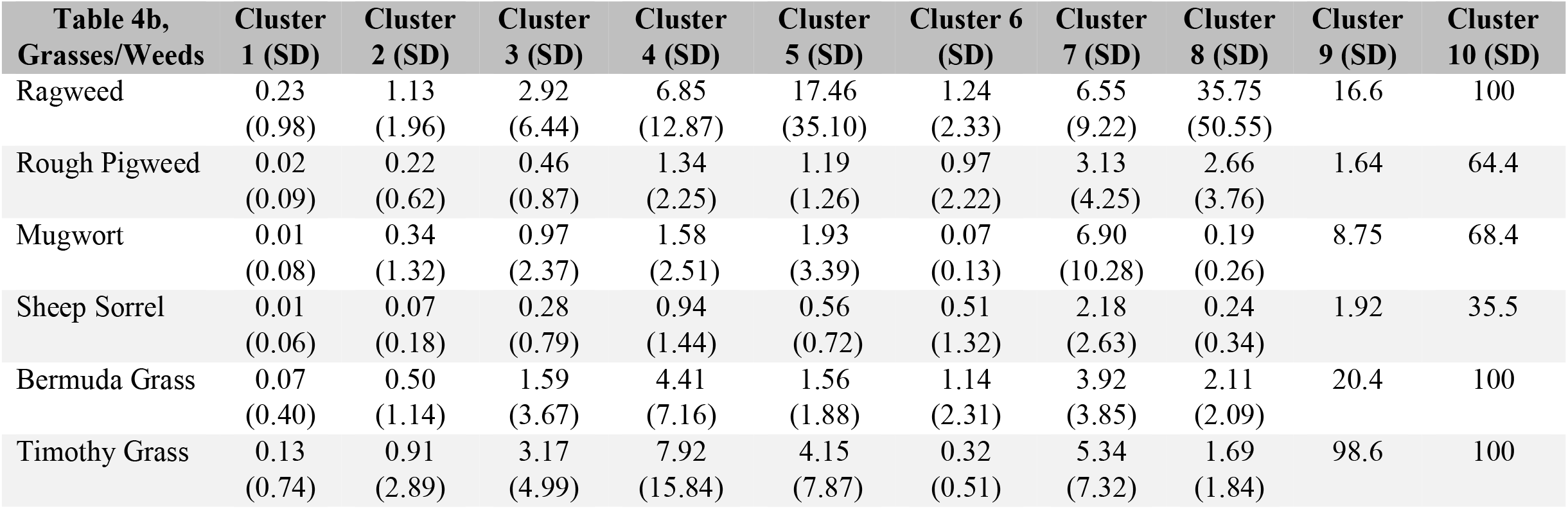
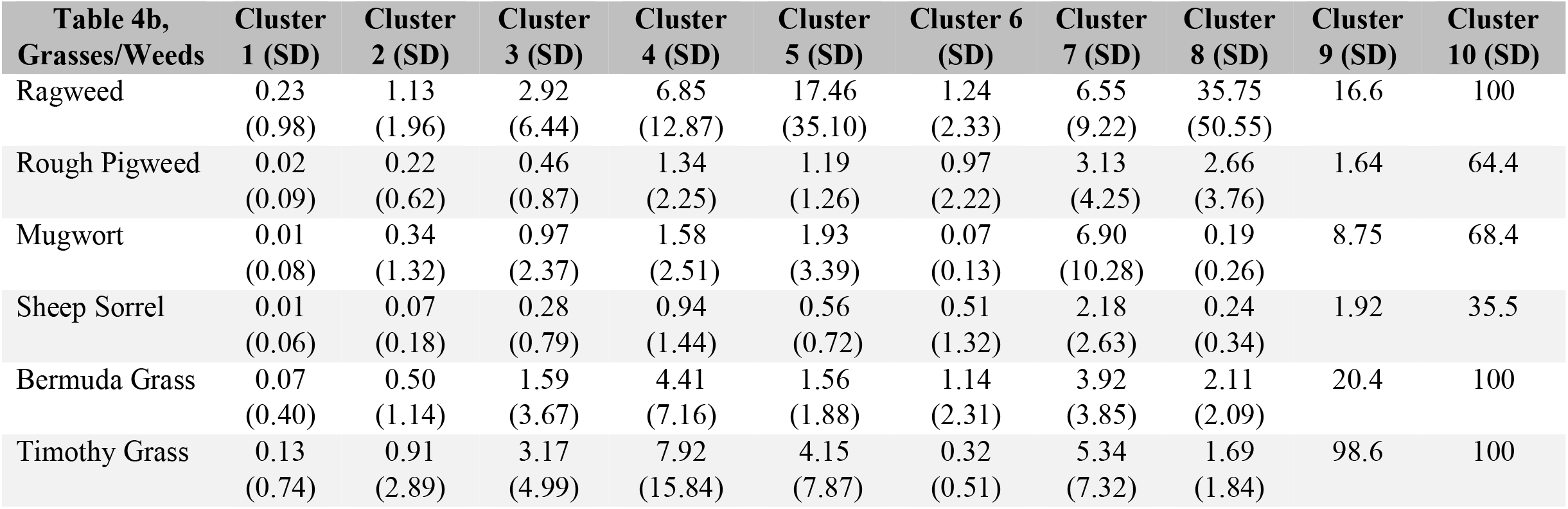
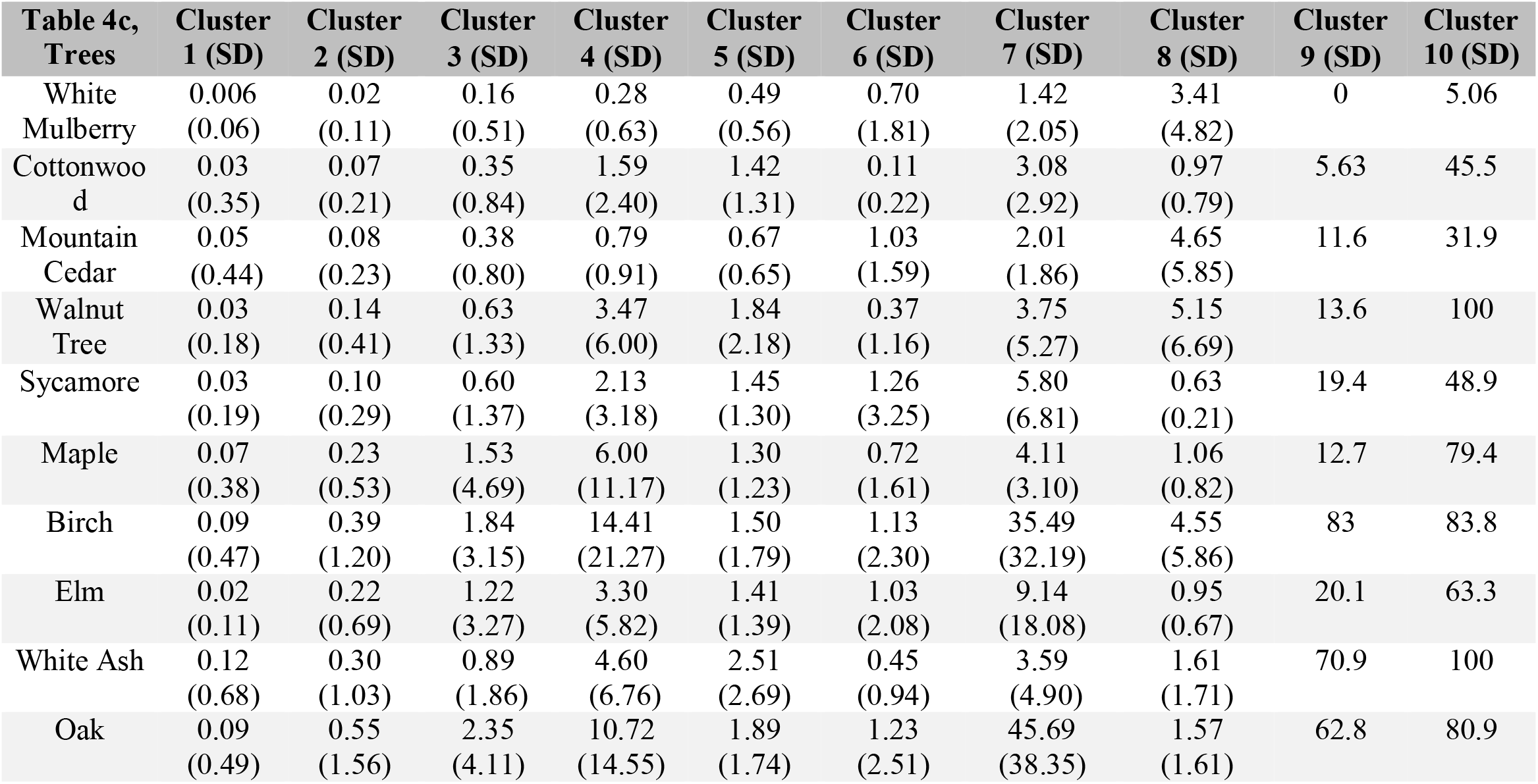
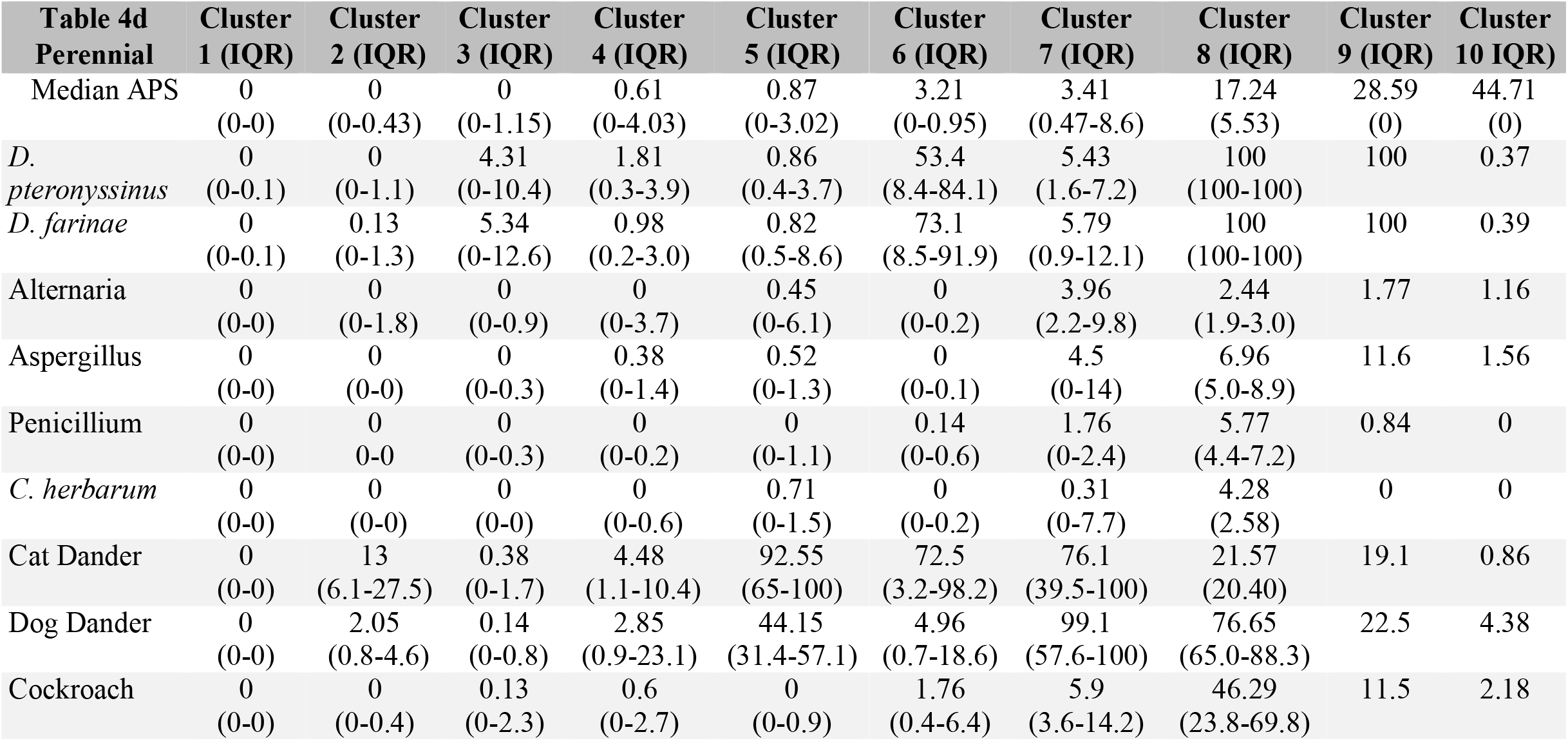
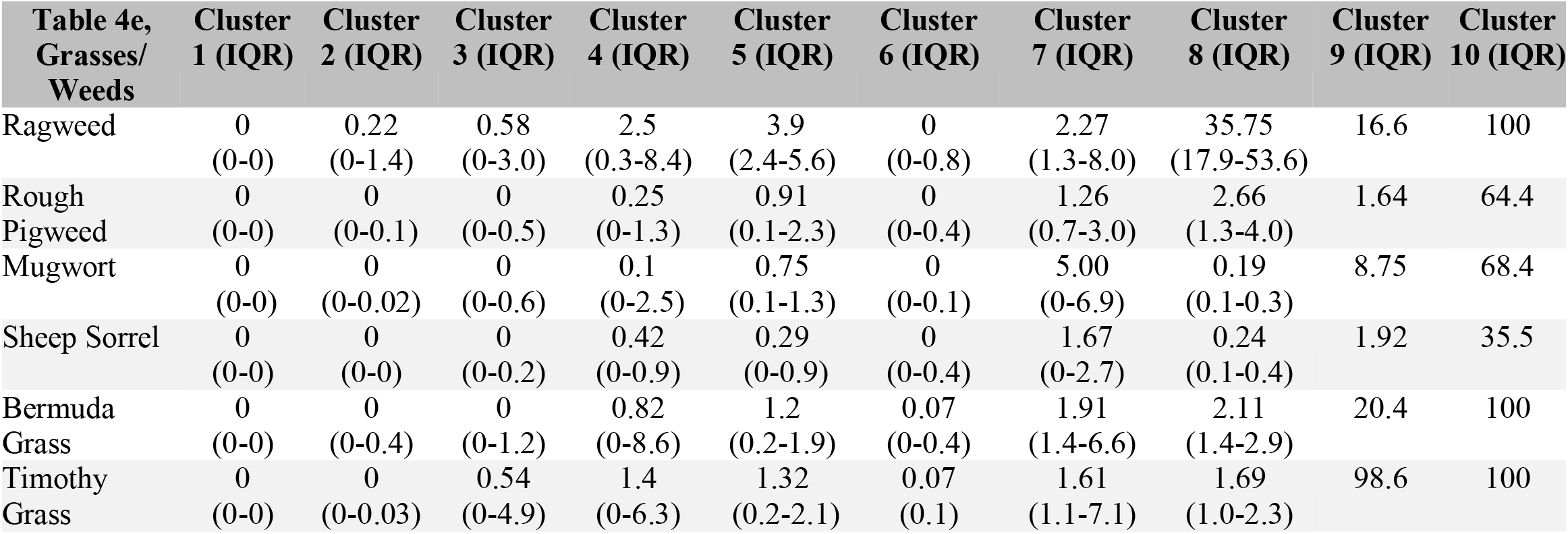
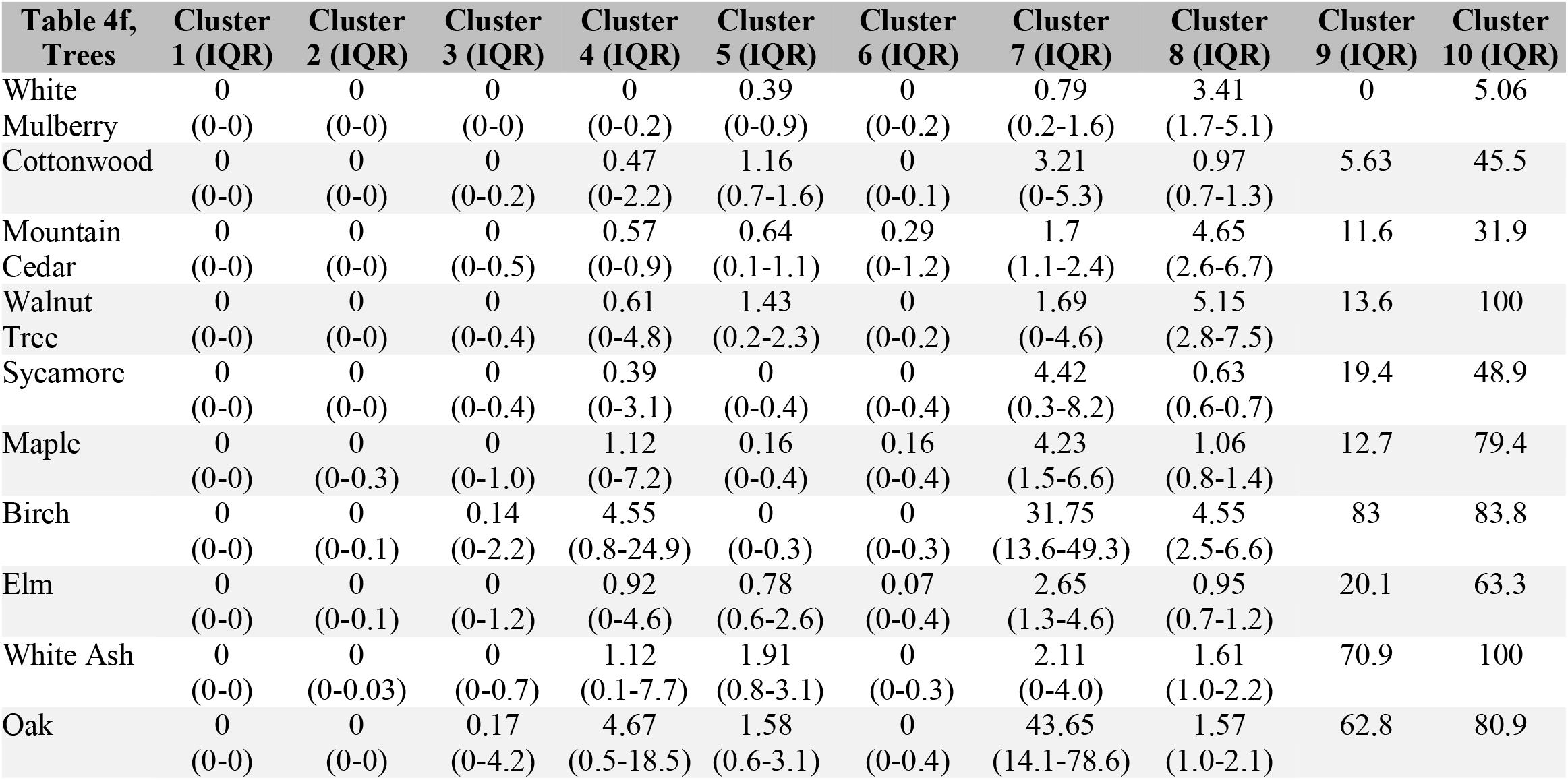
Sensitizations for each cluster (a-c); means and standard deviations (in parenthesis) of each cluster’s Allergic Poly-Sensitization (APS) score, calculated by the average specific Immunoglobulin E (IgE) for each patient in the cluster, and within cluster specific IgE. Median and interquartile range (in parenthesis) (d-f).

Cluster 8 only had two patients with very high sensitizations to perennial allergens, and one of the largest readings to cockroach. Cluster 9 and cluster 10 were both single patients. Surprisingly, these two patients with the highest APS did not have the highest total IgE (3,214 IU/ml and 2,750 IU/ml, respectively, supplementary Tabble 4), nor did the subject in cluster 9 have a particularly high PBEC (244/ul; cluster 10 patient not obtained). The patient in cluster 9 had an APS of 28.59 IU/ml, driven by reaching the upper limit of detection for *D. pteronyssinus* and *D. farinae*, nearly so for timothy grass and severe sensitizations to Bermuda grass, sycamore, oak, and white ash tree. The cluster 10 patient, despite sensitizations to *D. pteronyssinus, D. farinae, C. herbarum*, and *P. chrysogenum*, comparable to cluster 1 patients, displayed the upper limit for detection for five allergens (combined mean=44.71 IU/ml), approximately 400-fold higher than the average cluster 1 patient. Moreover, the patient’s highest sensitizations exceeded all other database patients for 12 of the 25 allergens.

### Seasonal vs Perennial Divergence of Clusters

Factor analysis of the sensitizations were plotted on a correlation circle. Two groups of allergens emerged as orthogonal to each other. (Fig 1d). This indicated that within the groups, the allergens were strongly and positively correlated and relatively uncorrelated to the allergens in the other group. The first principal component (horizontal axis) of the correlation circle explained 47.2% of the variance and was comprised of seasonal allergens (Fig 1b). The second principal component (vertical axis) explained 14.4% of the data variance and was comprised of perennial allergens (Fig 1c). Interestingly, sensitization to birch, oak, and white mulberry trees appeared to violate this trend.

**FIGURE 1:**
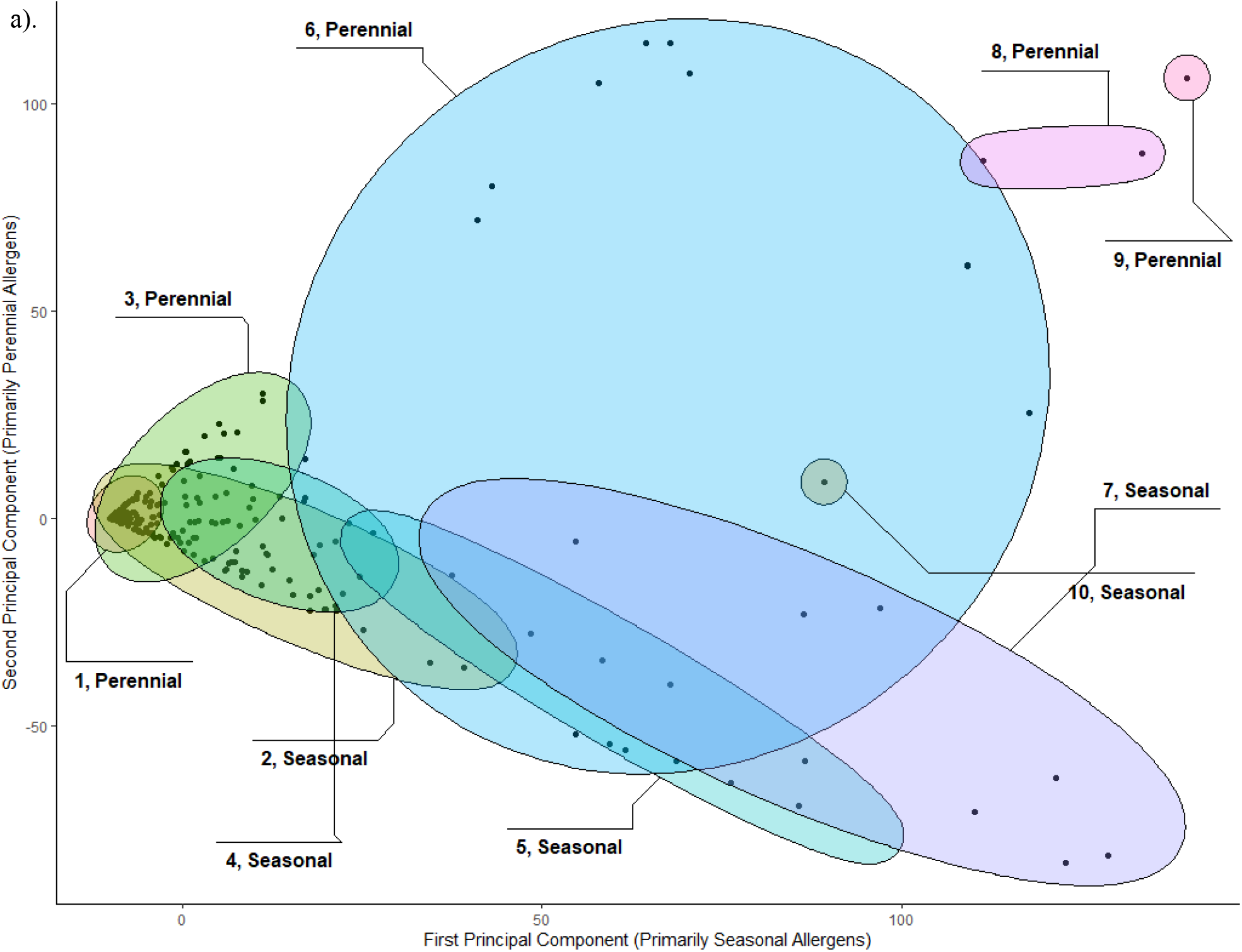
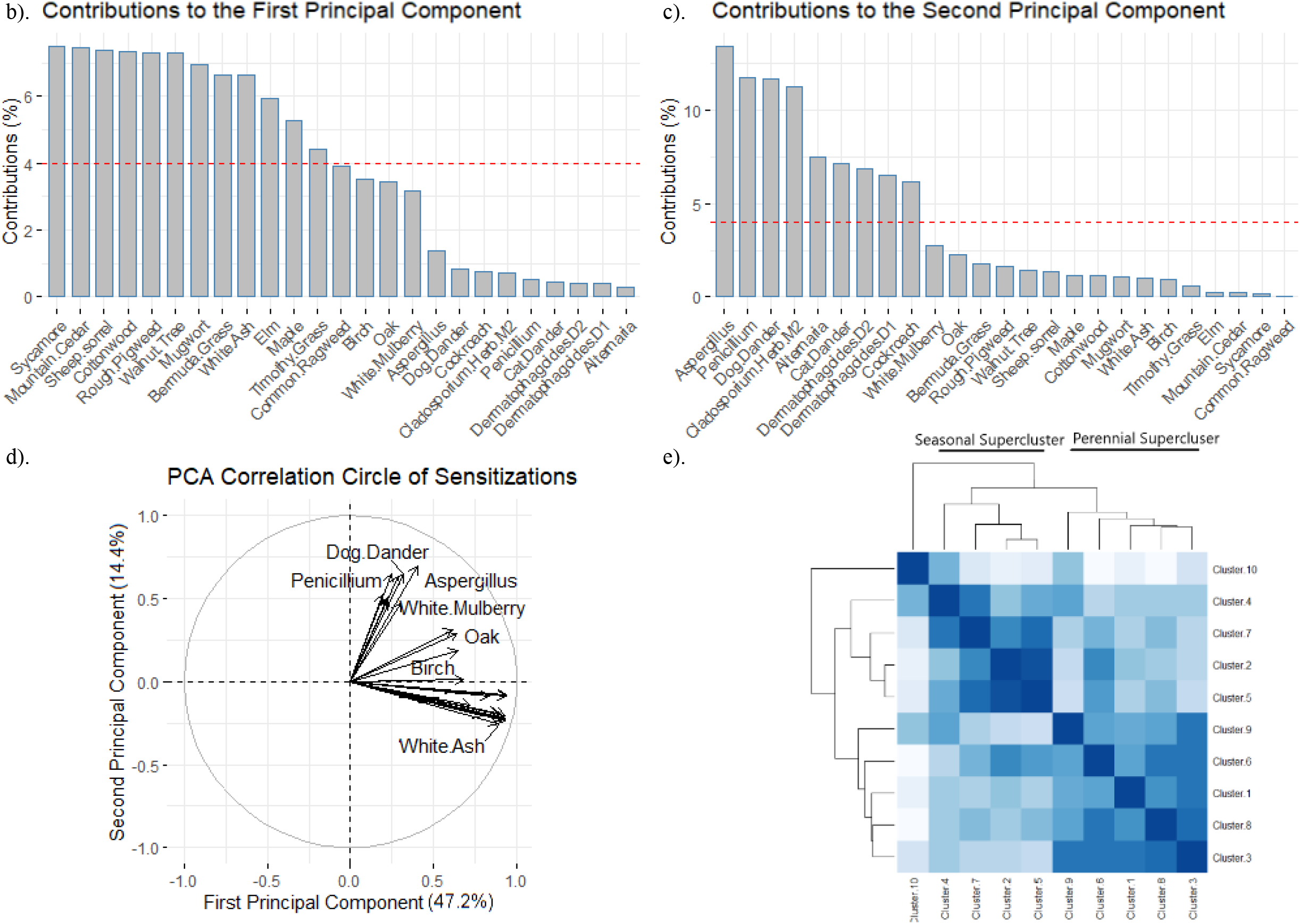
Cluster Correlations. The ten clusters generated by the Dirichlet model are shown and labeled with their cluster number, **1a**. Clusters are distinguished by both number of sensitizations and high specific Immunoglobulin E levels. The x-axis is the first principal component composed of primarily seasonal allergens detailed in **1b**, and the y-axis is the second principal component composed of primarily perennial allergens detailed in **1c**; red dashed lines indicate expected value if seasonal or perennial contributions were uniform. Principal Component Analysis (PCA) correlation circle with percent contributions to principal components 1 and 2 is shown in **1d**. Between-cluster correlation matrix of sensitizations ordered by hierarchical clustering, is shown in **1e**; darker shades indicate higher correlations.

Knowing these group separations, we hypothesized this would translate into polarized clusters that were individually homogenous for either perennial or seasonal allergens. To examine this possibility, we created a correlation matrix of the clusters and identified three “superclusters” (demonstrated in figure 1e). The first was comprised of clusters 1, 3, 6, 8, and 9; the second was made up of clusters 2, 4, 5, and 7; the third was just the single patient in cluster 10. The sensitizations in the first supercluster tended to be low for the perennial allergens but progressively higher to seasonal allergens. In contrast, the second supercluster showed progressively higher select perennial allergen sensitizations (i.e., mold, dust, cockroach) while comparatively modest sensitizations to seasonal allergens (Fig 1e). While the patient in cluster 10 was in their own supercluster, this person was qualitatively like the first supercluster (i.e. seasonal > perennial), though seasonal specific IgEs were far greater. Sensitizations to cat and dog appeared comparable in all three superclusters and non-compartmentalized.

### Significant Demographic Trends with Higher APS

Regression analysis was performed to assess the relationship between APS and common clinical markers of asthma. Findings are reported in supplementary tabble 3 and plots are reported in Figure 3, each sequential point in the plot (left to right) represents clusters 1 through 10. A simple linear regression evaluated the relationship between age and APS cluster assignment (Fig 2a). There was a significant negative correlation between APS and age (p=3.98e-05). No relationship between APS and BMI was predicted by simple linear regression (p=.197, Fig 2b). We did not find any significant differences when comparing clusters for differences in gender, race, or smoking history.

**FIGURE 2:**
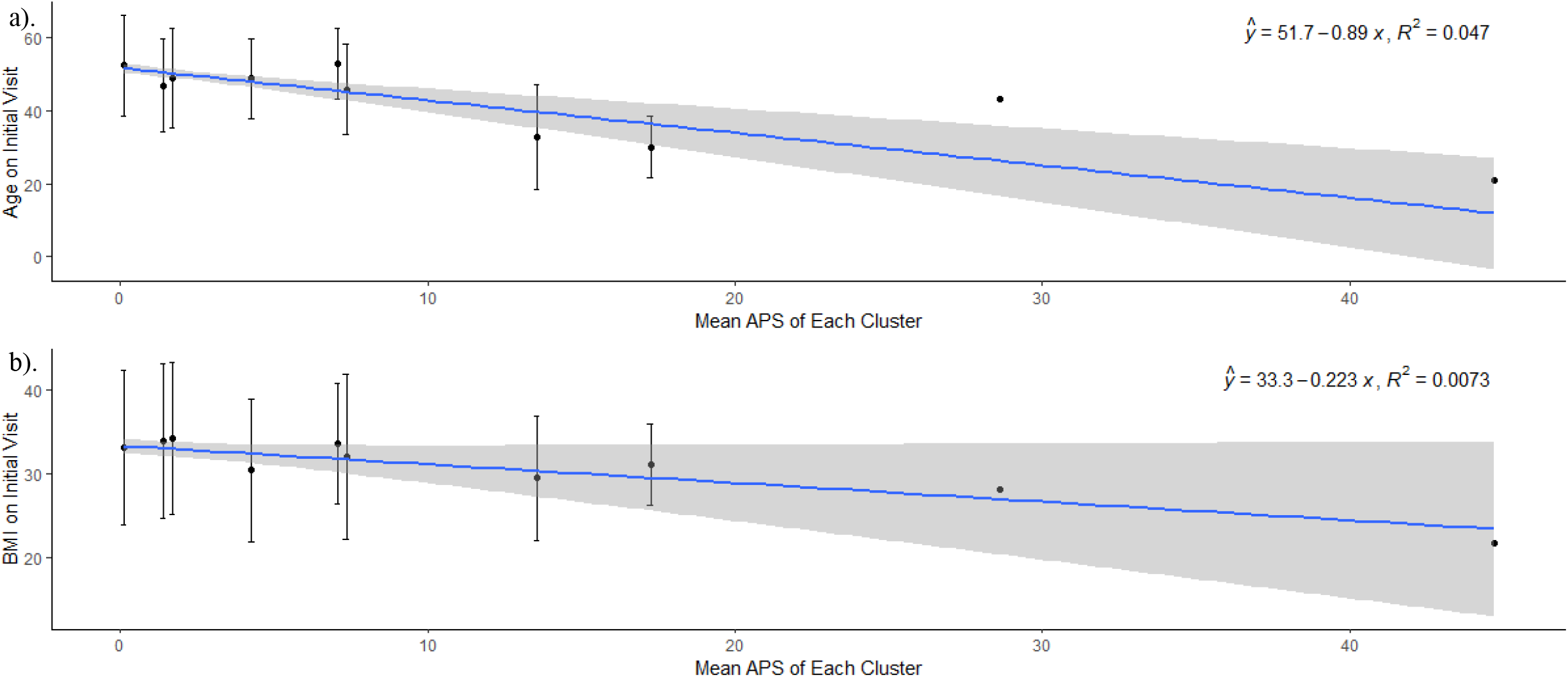
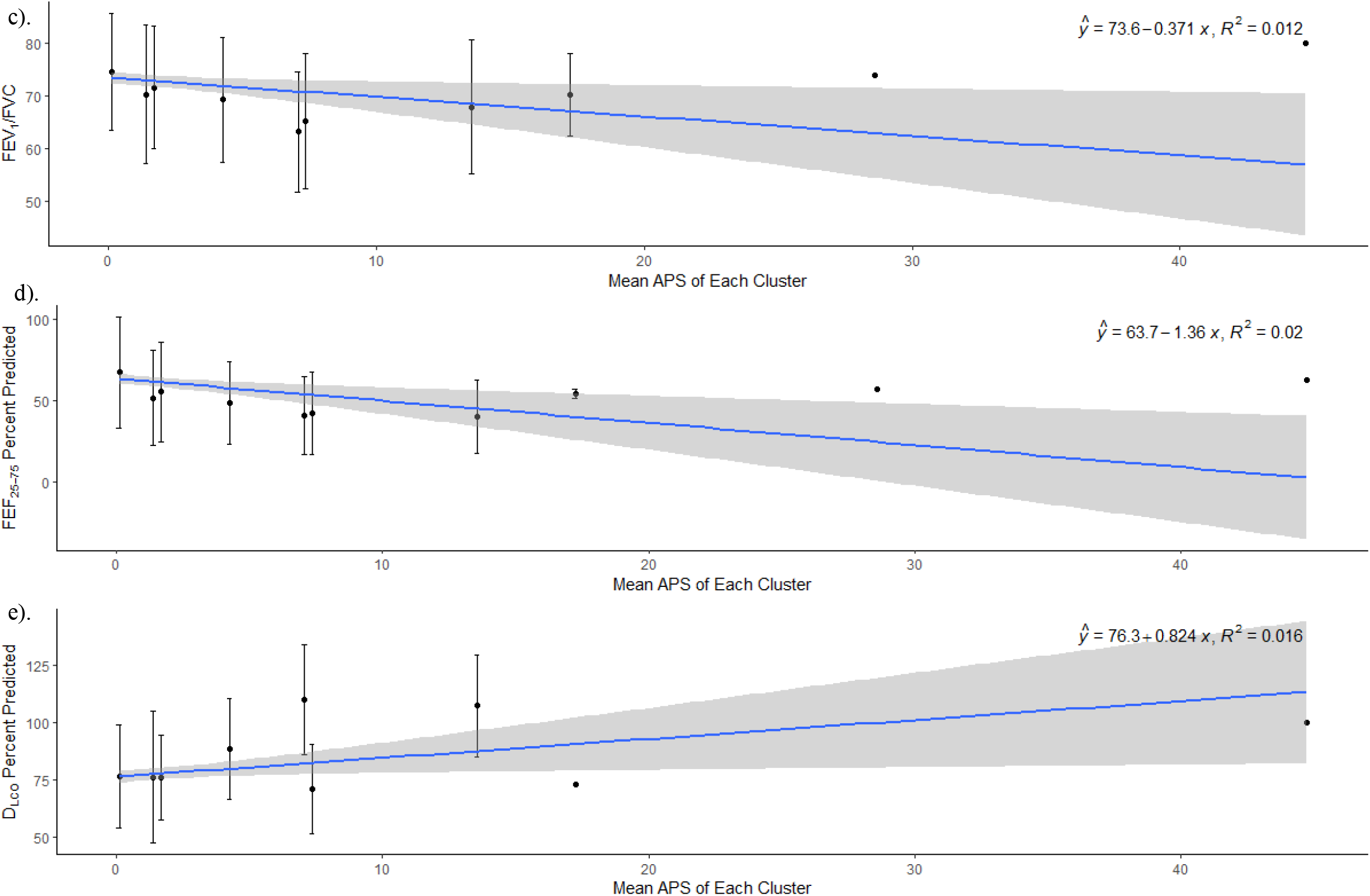
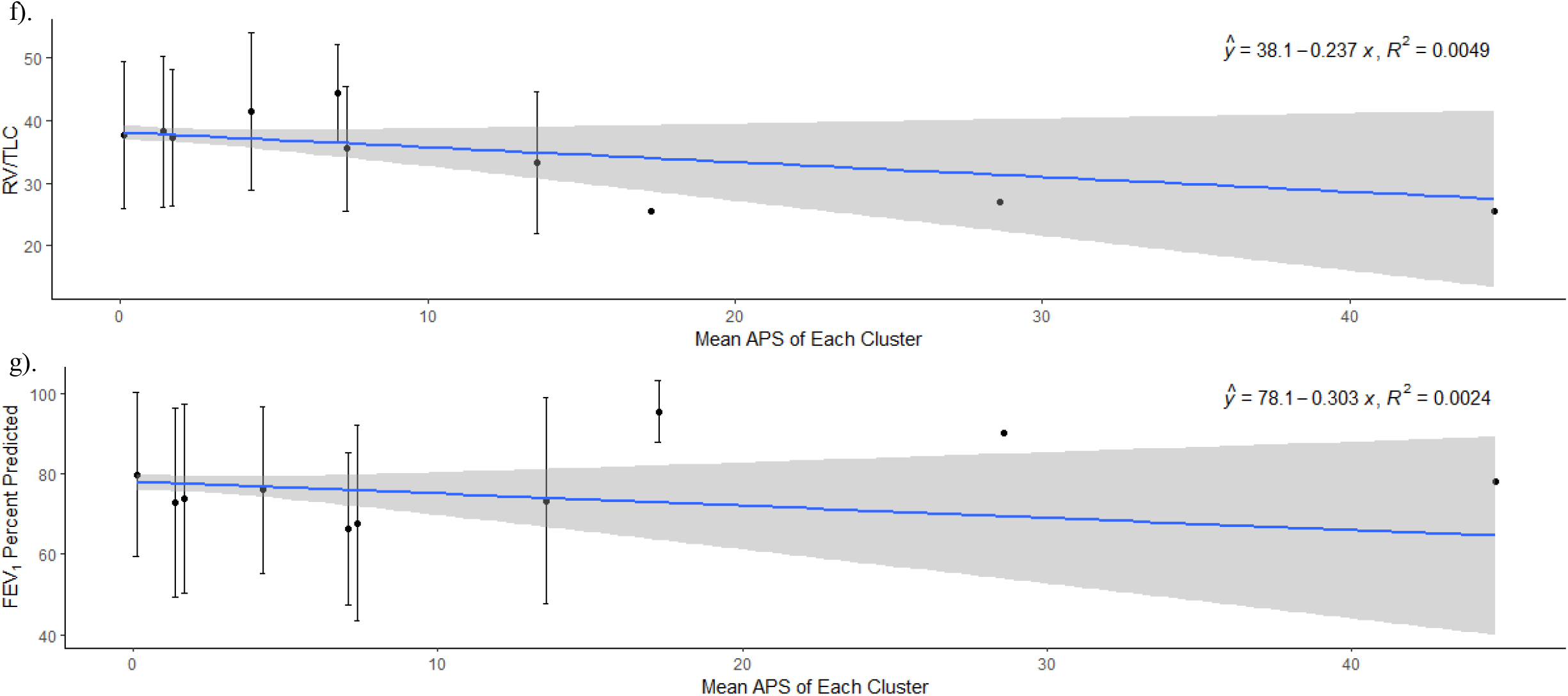
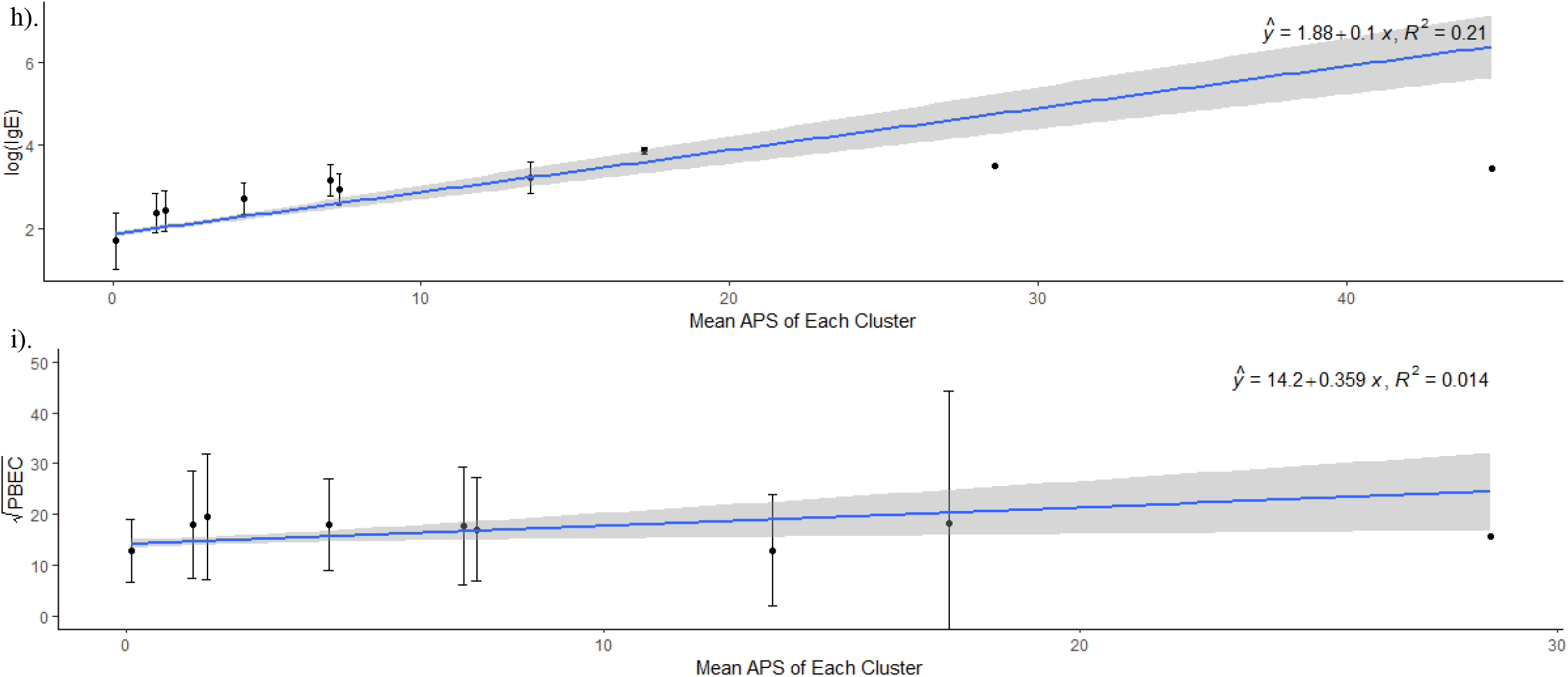
Linear Regression Plots. (a-i) APS for each sequential cluster is plotted left to right (clusters 1 through 10), along the x-axes by their APS score and clinical variables are plotted along y-axes. APS is calculated by average sensitization level to all allergens for all patients in the cluster. Points are mean ± SD. Overlaid is the best fit (blue line) from regression analysis and 95% confidence interval of regression line.

### APS Correlated with More Severe Clinical Markers of Allergic Asthma

Overall, higher APS was associated with higher obstruction on PFTs (Fig 2c-g). There was a statistically significant negative correlation between APS and FEV_1_/FVC (p=.01, Fig 2c) as well as FEF_25-75_ percent predicted (p=.002, Fig 2d). There was a statistically significant positive correlation between APS and D_LCO_ percent predicted (p=.01, Fig 2e). There were no statistically significant correlations between APS and RV/TLC (p=.14, Fig 2f) or FEV_1_ percent predicted (p=.26, Fig 2g).

The data for serum IgE and PBEC were highly non-linear and therefore required transformation prior to linear regression analysis. The data for serum IgE was log-transformed prior to regression analysis (not shown). There was a small percentage of patients (<5%) who had 0 to “<50/ul” PBEC recorded. Therefore, the square root was used to best achieve linearity. A significant regression equation was found between APS and the logarithm of serum IgE (p<2.2e-16, Fig 2h) and between APS and the square root of PBEC (p=.01, Fig 2i).

### Linear Regression After Outlier Removal

After plotting the regression analysis, we considered the possibility that the four patients in clusters 8, 9, and 10 were acting as outliers and affecting the results. To assess this, we repeated simple linear regression after removing these patients. The repeat analysis led to a significant regression equation between FEV_1_ percent predicted and APS (p=.01, supp tabble 3). The regression equations after removal of the outliers for the above variables were unchanged with respect to statistical significance.

## DISCUSSION

Our study population consisted of patients with specialist diagnosed, moderate to severe asthma mostly from zip codes below the federal poverty line. The majority were racial minorities (Table 3). Vulnerable patients in low resource areas have well-documented deficiencies in health outcomes^41^. This underscores the need for tools to improve care that are low cost and accessible to all respiratory practitioners with a routine blood draw^42^. Previous cluster analyses have relied in-part on clinical markers not available in many practitioner’s offices including exhaled nitric oxide and sputum eosinophilia vs neutrophilia^4, 5, 43^. Furthermore, no previous cluster analysis (Table 1) has used state-of-the-art quantitative allergen identification methods to examine if clusters matched for allergens, their levels of sensitization (0.1 to 100 IU/ml) and expressions of co-sensitizations would yield meaningful and clinically actionable insights. Atopy, when considered, was a uniform binary metric, failing to capture potential heterogeneity^44^. We turned to Dirichlet Process mixture modeling^29^, a well-established machine learning approach to impartially analyze our dataset of Zone 1 Northeastern US ImmunoCAP® 25 aeroallergen profiles. In multiple studies ImmunoCAP® assays are shown to be highly reproducible and accurate^20, 21^. We probed APS clusters and found retrospective assignments to predict several important clinical markers of asthma.

Unlike other clustering algorithms, mixture modeling used here is a probabilistic approach and allows for the prospective assignment of new patients with novel APS patterns. Although we applied modeling to the Zone 1 Northeastern US ImmunoCAP®, the collaborative development of mixture models at the population level for each ImmunoCAP® zone could be used to translate previously transactional sensitization reports into more clinically applicable information. We also aimed to have our approach serve as a model for future work where APS clustering could enhance comprehensive databases and be available in any connected location. APS clustering may lead to further understanding of how specific sensitization patterns are influenced by additional asthma risk factors (e.g., genetics, microbiome). Insights into pharmacologic outcomes, especially for biologic therapies^44^ may be gained if examined in relation to APS clusters. Lastly, recognition that atopy is not a single metric, APS as a biomarker may prove important as a standardized variable in future asthma cluster analyses (table 1) and could enhance databases for understanding other respiratory conditions (e.g., asthma-COPD overlap)^45^.

We used Dirichlet Process mixture models, which are an example of Bayesian non-parametric mixture models. Cluster number, size, shape, and cluster assignment were determined by a VB approach, allowing these to be independent, unbiased, algorithm-driven, and determined “automatically”. Non-parametric in this context means these models can have unbounded, growing, or infinite number of parameters (cluster number or complexity). Therefore, the Dirichlet Process has applications in many fields^46, 47^ where potentially new observations represent a novel cluster rather than a prior one. With just 25 aeroallergens in zone 1 ImmunoCAP® assays where specific IgEs range from 0.10 to 100 IU/ml, there are almost a googol (10^100^) possible profiles. To account for the possibility our datasets are incomplete, we turned to DPMMs, though this along with clustering in four principal components led to a large low APS cluster (cluster 1) and several small, high APS clusters. We acknowledge that clustering in fewer dimensions led to fewer and larger clusters, but also lost the ability to identify rare profiles and seasonal vs perennial trends. Our expectation is that incorporating multifold more patients will lead to these high APS, intensely atopic clusters expanding.

Overall, cluster analysis for APS yielded several notable clinical findings. We show clusters with higher specific IgE presented to the asthma subspeciality clinic at a younger age. Another finding was the strong positive correlation between APS and the Th2 biomarkers, total IgE and the PBEC. Finally, there was a significant negative correlation between the FEV_1_/FVC ratio and FEF_25-75_ percent predicted with a higher APS score. Surprisingly, the correlation between APS score and FEV_1_ percent predicted was not statistically significant, however, we found by elimination, this unexpected result was influenced by the four patients in clusters 8 through 10.

Hierarchical clustering indicated three major distinguishable classes of APS superclusters (Fig 2), which may represent divergent manifestations of the poly-atopic march. The first was made up of patients who tended to have progressively increasing perennial sensitization, while the other tended to have progressively increasing seasonal sensitization. Interestingly, the hierarchical clustering algorithm segregated the one cluster 10 patient profile as representing a third supercluster, though we tend to favor the view that they belong to the seasonal supercluster (see allergen profile, table 4).

Cluster analyses to-date have been heavily weighted on Euro-American Caucasian populations and not urban Black and Hispanic (Latino) patients. However, both a strength and limitation of our study is our demographic make-up. The limited diversity, particularly the low number of Caucasians and the preponderance of middle-aged, female, obese Afro-Americans from high poverty zip codes, should be considered when extending our findings to the moderate to severe asthma populations at large. The inclusion of diverse populations, particularly Euro-American Caucasians, is needed in future studies of APS.

To date, clustering approaches in asthma have not considered which allergens are positive, numbers of positive allergens, potential interactions of allergens affecting phenotypes, and levels of sensitization. Our work represents the first attempt to define poly-sensitization and how it may affect and refine asthma cluster analyses. Our clustering approach may lead to intuitive clinical questions including if treatment can be predictably tailored to each cluster, if the seasonal supercluster patients are indeed at an increased risk for seasonal exacerbations, and many others. Our studies show significant heterogeneity in the poly-atopic population, which should be considered in future approaches to clustering. Understanding the importance of APS will also help physicians understand the spectrum of sensitivities that drive patients’ symptoms, the poly-atopic march, and possibly optimize treatment strategies.

## Data Availability

All data produced in the present study are available upon reasonable request to the authors

## Acknowledgements

We thank the Margaret Wolf Memorial Pulmonary Research Fund for providing financial support for this research. The authors thank Dr. Phillip E. Silkoff for review of the manuscript.

## ABBREVIATIONS

APS: Allergic Poly-Sensitization
ACO: Asthma-COPD overlap
BMI: Body Mass Index
CBC: Complete Blood Count
CI: Confidence Interval
COPD: Chronic Obstructive Pulmonary Disease
D_LCO_: Diffusion Capacity of the Lungs for Carbon Monoxide
DPMM: Dirichlet Process Mixture Model
D1: *Dermatophagoides pteronyssinus*
D2: *Dermatophagoides farinae*
FEF_25-75_: Forced expiratory flow at 25–75% of forced vital capacity
FEV_1_: Volume of air forcibly expired in 1 second
FEV_1_/FVC: Ratio of air forcibly expired in 1 second to the forced vital capacity
FVC: Forced Vital Capacity
IgE: Immunoglobulin E
IQR: Interquartile Range
IU/ml: International units per milliliter
PCA: Principal Component Analysis
PFT: Pulmonary Function Test
RV: Residual Volume
RV/TLC: Ratio of residual volume to total lung capacity
SARP: Severe Asthma Research Program
SD: Standard Deviation
TLC: Total Lung Capacity
VB: Variational Bayes

## Notes

**Conflicts of Interest:** No conflicts of interest to disclose

### Competing Interest Statement

The authors have declared no competing interest.

### Author Declarations

The research was approved by the Institutional Review Board of the Drexel University College of Medicine who gave ethical approval for this work in an expedited review.

## REFERENCES

1. Dharmage SC, Perret JL, Custovic A. Epidemiology of Asthma in Children and Adults. Front Pediatr. 2019;7:246. doi:10.3389/fped.2019.00246

2. Agache I, Akdis CA. Endotypes of allergic diseases and asthma: An important step in building blocks for the future of precision medicine. Allergol Int. Jul 2016;65(3):243–52. doi:10.1016/j.alit.2016.04.011

3. Kaur R, Chupp G. Phenotypes and endotypes of adult asthma: Moving toward precision medicine. J Allergy Clin Immunol. 07 2019;144(1):1–12. doi:10.1016/j.jaci.2019.05.031

4. Haldar P, Pavord ID, Shaw DE, et al. Cluster analysis and clinical asthma phenotypes. Am J Respir Crit Care Med. Aug 01 2008;178(3):218–224. doi:10.1164/rccm.200711-1754OC

5. Moore WC, Meyers DA, Wenzel SE, et al. Identification of asthma phenotypes using cluster analysis in the Severe Asthma Research Program. Am J Respir Crit Care Med. Feb 15 2010;181(4):315–23. doi:10.1164/rccm.200906-0896OC

6. Amelink M, de Groot JC, de Nijs SB, et al. Severe adult-onset asthma: A distinct phenotype. J Allergy Clin Immunol. Aug 2013;132(2):336–41. doi:10.1016/j.jaci.2013.04.052

7. Kim TB, Jang AS, Kwon HS, et al. Identification of asthma clusters in two independent Korean adult asthma cohorts. Eur Respir J. Jun 2013;41(6):1308–14. doi:10.1183/09031936.00100811

8. Silkoff PE, Strambu I, Laviolette M, et al. Asthma characteristics and biomarkers from the Airways Disease Endotyping for Personalized Therapeutics (ADEPT) longitudinal profiling study. Respir Res. Nov 17 2015;16:142. doi:10.1186/s12931-015-0299-y

9. Fitzpatrick AM, Moore WC. Severe Asthma Phenotypes - How Should They Guide Evaluation and Treatment? J Allergy Clin Immunol Pract. 2017 Jul - Aug 2017;5(4):901–908. doi:10.1016/j.jaip.2017.05.015

10. Schatz M, Hsu JW, Zeiger RS, et al. Phenotypes determined by cluster analysis in severe or difficult-to-treat asthma. J Allergy Clin Immunol. Jun 2014;133(6):1549–56. doi:10.1016/j.jaci.2013.10.006

11. Newby C, Heaney LG, Menzies-Gow A, et al. Statistical cluster analysis of the British Thoracic Society Severe Refractory Asthma Registry: clinical outcomes and phenotype stability. PLoS One. 2014;9(7):e102987. doi:10.1371/journal.pone.0102987

12. Zaihra T, Walsh CJ, Ahmed S, et al. Phenotyping of difficult asthma using longitudinal physiological and biomarker measurements reveals significant differences in stability between clusters. BMC Pulm Med. May 10 2016;16(1):74. doi:10.1186/s12890-016-0232-2

13. Silkoff PE, Moore WC, Sterk PJ. Three Major Efforts to Phenotype Asthma: Severe Asthma Research Program, Asthma Disease Endotyping for Personalized Therapeutics, and Unbiased Biomarkers for the Prediction of Respiratory Disease Outcome. Clin Chest Med. 03 2019;40(1):13–28. doi:10.1016/j.ccm.2018.10.016

14. Deng K, Zhang X, Liu Y, et al. Heterogeneity of Paucigranulocytic Asthma: A Prospective Cohort Study with Hierarchical Cluster Analysis. J Allergy Clin Immunol Pract. 06 2021;9(6):2344–2355. doi:10.1016/j.jaip.2021.01.004

15. Lefaudeux D, De Meulder B, Loza MJ, et al. U-BIOPRED clinical adult asthma clusters linked to a subset of sputum omics. J Allergy Clin Immunol. Jun 2017;139(6):1797–1807. doi:10.1016/j.jaci.2016.08.048

16. Platts-Mills TA. The role of immunoglobulin E in allergy and asthma. Am J Respir Crit Care Med. Oct 15 2001;164(8 Pt 2):S1–5. doi:10.1164/ajrccm.164.supplement_1.2103024

17. Djukanovic R, Wilson SJ, Kraft M, et al. Effects of treatment with anti-immunoglobulin E antibody omalizumab on airway inflammation in allergic asthma. Am J Respir Crit Care Med. Sep 15 2004;170(6):583–93. doi:10.1164/rccm.200312-1651OC

18. Migueres M, Dávila I, Frati F, et al. Types of sensitization to aeroallergens: definitions, prevalences and impact on the diagnosis and treatment of allergic respiratory disease. Clin Transl Allergy. 2014;4:16. doi:10.1186/2045-7022-4-16

19. Ciprandi G, Cirillo I. Monosensitization and polysensitization in allergic rhinitis. Eur J Intern Med. Dec 2011;22(6):e75–9. doi:10.1016/j.ejim.2011.05.009

20. Szeinbach SL, Barnes JH, Sullivan TJ, Williams PB. Precision and accuracy of commercial laboratories’ ability to classify positive and/or negative allergen-specific IgE results. Ann Allergy Asthma Immunol. 2001 Apr;86(4):373–81. doi: 10.1016/S1081-1206(10)62481-7. PMID: 11345278.

21. Leimgruber A, Mosimann B, Claeys M, et al. Clinical evaluation of a new in-vitro assay for specific IgE, the immuno CAP System. Clin Exp Allergy 1991;21:127–131.

22. Schulman ES, Pohlig C. Rationale for specific allergen testing of patients with asthma in the clinical pulmonary office setting. Chest. Jan 2015;147(1):251–258. doi:10.1378/chest.12-0072

23. Chung KF, Wenzel SE, Brozek JL, Bush A, Castro M, Sterk PJ, Adcock IM, Bateman ED, Bel EH, Bleecker ER, Boulet LP, Brightling C, Chanez P, Dahlen SE, Djukanovic R, Frey U, Gaga M, Gibson P, Hamid Q, Jajour NN, Mauad T, Sorkness RL, Teague WG. International ERS/ATS guidelines on definition, evaluation and treatment of severe asthma. Eur Respir J. 2014 Feb;43(2):343–73. doi: 10.1183/09031936.00202013. Epub 2013 Dec 12.

24. Global Strategy for Asthma Management and Prevention. Fontana, Global Initiative for Asthma, 2021. https://ginasthma.org/reports/ (Last accessed February 20, 2022)

25. González Burchard E, Borrell LN. Need for Racial and Ethnic Diversity in Asthma Precision Medicine. N Engl J Med. 12 09 2021;385(24):2297–2298. doi:10.1056/NEJMe2114944

26. Semega J, Kollar M, Shrider EA, Creamer JF. Income and Poverty in the United States: 2019. U.S. Census Bureau, Current Population Reports, 2020; 60–270.

27. Data Access | City Health Dashboard. Retrieved October 5, 2020, from https://www.cityhealthdashboard.com/data-access

28. Cattell RB. The Scree Test For The Number Of Factors. Multivariate Behav Res. Apr x01 1966;1(2):245–76. doi:10.1207/s15327906mbr0102_10

29. Bishop CM. Pattern recognition. Machine learning. 2006 Feb;128(9).

30. Pedregosa F, Varoquaux G, Gramfort A, et al. Scikit-learn: Machine learning in Python. J Mach Learn Res. 2011; 12: 2825–2830

31. Blei DM, Jordan MI. Variational inference for Dirichlet process mixtures. Bayesian Anal. 2006; 1(1) 121 –143

32. Attias H. A Variational Bayesian Framework for Graphical Models. Adv Neural Inf Process Syst. 2000; 12.

33. Pitman J. Combinatorial Stochastic Process. 2002 https://www.stat.berkeley.edu/~pitman/621.pdf

34. Abdi H, Williams LJ. Principal components analysis. Wiley Interdiscip. Rev. Comput. Stat. 2010; 2: 433–450

35. Wei T, Simko V, Levy M, et al. “Package ‘corrplot’.” Statistician 2017; 56.316: e24.

36. Kassambara A, Mundt F. Package “factoextra” Extract and Visualize the Results of Multivariate Data Analyses 2017.

37. Lê S, Josse J, Rennes A, Husson F. FactoMineR: An R Package for Multivariate Analysis. J Stat Softw, 2015; 25

38. R: The R Project for Statistical Computing. 2013; https://www.r-project.org/

39. Wickham H. ggplot2. Wiley Interdiscip. Rev. Comput. Stat. 2011; 3(2), 180–185.

40. Wickham H, Francois R, Henry L, Müller K. (2015). dplyr: A grammar of data manipulation. R package version 0.4, 3, p156.

41. (CDC) CfDCaP. Asthma prevalence and control characteristics by race/ethnicity--United States, 2002. MMWR Morb Mortal Wkly Rep. Feb 27 2004;53(7):145–8.

42. Press VG, Pappalardo AA, Conwell WD, Pincavage AT, Prochaska MH, Arora VM. Interventions to improve outcomes for minority adults with asthma: a systematic review. J Gen Intern Med. Aug 2012;27(8):1001–15. doi:10.1007/s11606-012-2058-9

43. Heaney LG, McGarvey LP. Personalized Medicine for Asthma and Chronic Obstructive Pulmonary Disease. Respiration. 2017;93(3):153–161. doi:10.1159/000455395

44. Dávila I, Campo P, Cimbollek S, et al. Cluster sub-analysis of patients with severe asthma who responded to omalizumab. J Investig Allergol Clin Immunol. Sep 28 2021:0. doi:10.18176/jiaci.0731

45. Leung C, Sin DD. Asthma-COPD Overlap: What Are the Important Questions? Chest. 2022 Feb;161(2):330–344. doi: 10.1016/j.chest.2021.09.036. Epub 2021 Oct 6. PMID: 34626594.

46. Favaro, S., Lijoi, A., Mena, R. H., & Prünster, I. (2009). Bayesian non_parametric inference for species variety with a two_parameter Poisson–Dirichlet process prior. Journal of the Royal Statistical Society: Series B (Statistical Methodology), 71(5), 993–1008.

47. Teh, Y.W. and Jordan, M.I., 2010. Hierarchical Bayesian nonparametric models with applications. Bayesian nonparametrics, 1, pp.158–207.

