## Supplementary files for "Cluster Analysis of Allergic Poly-Sensitizations in Urban Adults with Asthma"

### **SUPPLEMENTARY FIGURES**

**SUPPLEMENTARY TABLE 1:**

|  | **Allergens** |
| --- | --- |
| Perennial | *Dermatophagoides pteronyssinus* |
|  | *Dermatophagoides farinae* |
|  | *Alternaria alternata* |
|  | *Aspergillus fumigatus* |
|  | *Penicillium notatum* |
|  | *Cladosporium herbarum* |
|  | Cat Dander |
|  | Dog Dander |
|  | Cockroach |
| Grasses/Weeds | Ragweed |
|  | Rough Pigweed |
|  | Mugwort |
|  | Sheep Sorrel |
|  | Bermuda Grass |
|  | Timothy Grass |
| Trees | White Mulberry |
|  | Cottonwood |
|  | Mountain Cedar |
|  | Walnut Tree |
|  | Sycamore |
|  | Maple |
|  | Birch |
|  | Elm |
|  | White Ash |
|  | Oak |

**SUPPLEMENTARY TABLE 1:** Allergens tested in the Zone 1 ImmunoCAP

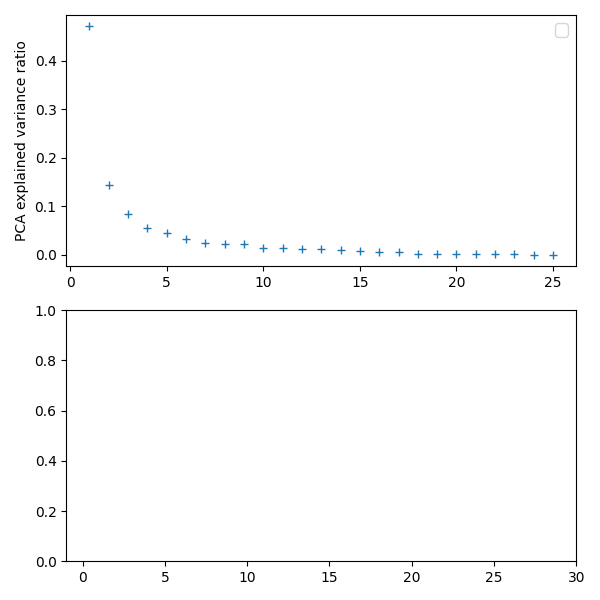

**SUPPLEMENTARY Figure 1**: Scree plot. Elbow was determined to be between 3-6, with 4 being chosen as the number of components for PCA dimensionality reduction.

**SUPPLEMENTARY TABLE 2:**

|  | **Cluster 1** | **Cluster 2** | **Cluster 3** | **Cluster 4** | **Cluster**  **5** | **Cluster**  **6** | **Cluster 7** | **Cluster**  **8** | **Cluster 9** | **Cluster 10** |
| --- | --- | --- | --- | --- | --- | --- | --- | --- | --- | --- |
| All allergens | 1  (0-3) | 6  (3.75-11) | 12  (5-17) | 16  (12-22) | 19.5  (16-21.5) | 10  (5-20) | 24.5 (24.5-25) | 22.5 (22.25-22.75) | 23 | 23 |
| Perennial | 1  (0-2) | 5  (3-6) | 5  (3-7.5) | 7  (5-10) | 8  (6.25-9.75) | 6  (4-10.25) | 10.5 (7-11) | 10.5 (10.25-10.75) | 10 | 9 |
| Grasses/weeds | 0  (0-1) | 1  (0-3) | 3  (1-5) | 4  (2-6) | 5.5  (2-6) | 2  (0-4.5) | 6  (3.75-6) | 4  (4-4) | 6 | 6 |
| Trees | 0  (0-0) | 0  (0-2) | 2  (0.5-5.5) | 5  (4-8) | 7  (4.75-7.75) | 1.5  (0-5.75) | 8  (3.75-8) | 8  (8-8) | 7 | 8 |

**SUPPLEMENTARY TABLE 2:** Median number of positive sensitizations for each cluster

**SUPPLEMENTARY TABLE 3:**

| **Variable** | **β_0_ (95% CI)** | **β_1_ (95% CI)** | **p-value** | **R^2^** |
| --- | --- | --- | --- | --- |
| Age | 51.7 (50.40, 53.01) | -0.89 (-1.24, -0.52) | 8.551e-07 | 0.0450 |
| BMI | 33.3 (32.41, 34.25) | -0.22 (-0.47, 0.02) | 0.07 | 0.005 |
| FEV_1_ | 78.1 (75.98, 80.218) | -0.30 (-1.42, 0.82) | 0.29 | 0.0002 |
| FEV_1_ after outlier removal | 78.8 (76.60, 81.00) | -1.09 (-1.97, -0.21) | 0.01 | 0.01 |
| FEV_1_/FVC | 73.6 (72.44, 74.76) | -0.37 (-0.68, -0.06) | 0.01 | 0.01 |
| FEF_25-75_ | 63.7 (60.42, 66.98) | -1.36 (-2.23, -0.49) | 0.002 | 0.02 |
| RV/TLC | 38.1 (36.87, 39.29) | -0.23 (-0.57, 0.09) | 0.14 | 0.004 |
| D_LCO_ | 76.3 (73.54, 79.04) | 0.82 (0.10, 1.54) | 0.02 | 0.01 |
| (PBEC)^1/2^ | 14.19 (13.31, 15.06) | 0.35 (0.08, 0.64) | 0.01 | 0.01 |
| log_10_(Total Serum IgE) | 1.88 (1.81, 1.94) | 0.10 (0.08, 0.12) | p<2.2e-16 | 0.20 |

**SUPPLEMENTARY TABLE 3:** Linear Regression Output. β_0_ (intercept) and β_1_ (slope) with 95% confidence interval (CI), p-value, and R^2^ are reported

**SUPPLEMENTARY TABLE 4a**

| **Clinical Data** | **Cluster**  **1** | **Cluster**  **2** | **Cluster**  **3** | **Cluster**  **4** | **Cluster 5** | **Cluster 6** | **Cluster 7** | **Cluster 8** | **Cluster 9** | **Cluster 10** |
| --- | --- | --- | --- | --- | --- | --- | --- | --- | --- | --- |
| FEV_1_ % (SD) | 79.8 (20.5) | 72.9 (23.4) | 73.8 (23.4) | 76.1 (20.7) | 66.3 (19.0) | 67.8 (24.3) | 73.3 (25.7) | 95.5 (7.78) | 90 | 78 |
| Missing, n (%) | 19 (6.0) | 3 (4.2) | 2 (8.3) | 6 (17.1) | 1 (8.3) | 0 | 2 (20) | 0 | 0 | 0 |
| FEV_1_/FVC (SD) | 74.7 (11.2) | 70.4 (13.3) | 71.6 (11.7) | 69.3 (11.6) | 63.2 (11.5) | 65.22 (12.9) | 68.0 (12.8) | 70.3 (7.9) | 74 | 80.1 |
| Missing, n (%) | 19 (6.0) | 2 (2.8) | 2 (9.3) | 5 (14.3) | 1 (8.3) | 0 | 2 (20) | 0 | 0 | 0 |
| FEF_25-75_ % (SD) | 67.4 (34.2) | 51.8 (29.4) | 55.6 (30.5) | 48.6 (25.2) | 40.8 (23.8) | 42.5 (25.5) | 40.3 (22.3) | 54.0 (2.82) | 57 | 63 |
| Missing, n (%) | 26 (8.2) | 3 (4.2) | 2 (8.3) | 6 (17.1) | 1 (8.3) | 0 | 2 (20) | 0 | 0 | 0 |
| RV/TLC  (SD) | 37.7 (11.8) | 38.2 (12.1) | 37.3 (10.9) | 41.5 (12.6) | 44.3 (7.7) | 35.5 (33.3) | 33.3 (11.4) | 25.4 | 27.1 | 25.5 |
| Missing, n (%) | 53 (16.8) | 9 (12.5) | 4 (16.7) | 8 (22.9) | 2 (16.7) | 0 | 2 (20) | 1 (50) | 0 | 0 |
| D_LCO_ % (SD) | 76.5 (22.3) | 76.1 (28.8) | 75.9 (18.3) | 88.5 (21.9) | 110.0 (24.0) | 71.1 (19.4) | 107.2 (22.3) | 73 | Not tested | 100 |
| Missing, n (%) | 129 (40.8) | 34 (47.2) | 4 (16.66) | 15 (42.9) | 6 (50) | 0 | 4 (40) | 1 (50) | 1 | 0 |

**SUPPLEMENTARY TABLE 4b**

| **Clinical Data** | **Cluster**  **1** | **Cluster**  **2** | **Cluster**  **3** | **Cluster**  **4** | **Cluster**  **5** | **Cluster**  **6** | **Cluster**  **7** | **Cluster**  **8** | **Cluster 9** | **Cluster 10** |
| --- | --- | --- | --- | --- | --- | --- | --- | --- | --- | --- |
| Serum IgE IU/ml (SD) | 190.1 (471.5) | 230.9 (451.0) | 923.13 (1561.4) | 790.5 (676.6) | 910.3 (586.4) | 408  (130.1) | 2614.8 (1720.4) | 7636.5 (1255.1) | 3214 | 2750 |
| Median  (IQR) | 44  (19-151) | 244.5  (116-528) | 250  (121-557) | 573  (291-956) | 1628  (845-3392) | 1073  (526-1211) | 1869  (1091-2752) | 7636  (7192-8080) | 3214 | 2750 |
| Missing, n  (%) | 11 (3.5) | 3 (4.2) | 1 (4.2) | 1 (2.9) | 0 | 2 (50) | 0 | 0 | 0 | 0 |
| PBEC  cells/ml (SD) | 203.3 (201.4) | 431.9 (556.3) | 200.1 (122.1) | 438.5 (435.3) | 149.2 (135.7) | 812.5 (997.2) | 461.9 (331.3) | 669.5 (946.8) | 244 | Missing |
| Median  (IQR) | 144.5  (91-258) | 183  (100-370) | 192  (100-370) | 342  (153-560) | 359  (145-746) | 265.5  (152-382) | 198  (31-442) | 669.5  (334-1004) | 244 | Missing |
| Missing, n  (%) | 34 (10.8) | 7 (9.72) | 1 (4.16) | 4 (11.4) | 2 (16.7) | 0 | 1 (10) | 0 | 0 | 1 (100) |

**SUPPLEMENTARY TABLE 4**: Within Cluster Clinical Markers of Asthma. (a) Mean and standard deviation are reported for each cluster; (b) serum Immunoglobulin E (IgE) and peripheral blood eosinophil count (PBEC) data were highly skewed, therefore for these median and interquartile range are also reported
